# Dancing Through Time: Cognitive Changes Over Six-Years of Community Dance in Parkinson’s Disease

**DOI:** 10.1101/2025.04.26.25326488

**Authors:** Simran Rooprai, Harsimran Dogra, Ashkan Karimi, Rafia Rafique, Emily D’Alessandro, Karolina Bearss, Sarah Robichaud, Rachel J. Bar, Joseph F.X. DeSouza

## Abstract

**Background:** Parkinson’s Disease (PD) is the most common neurodegenerative disorder after Alzheimer’s, and is characterized by motor and non-motor symptoms, including gait dysfunction and cognitive decline. Dance has emerged as a promising intervention for improving motor and non-motor symptoms in persons with PD (PwPD), yet long-term effects remain underexplored.

**Objective:** To assess changes in cognitive function and gait performance over six years among PwPD who participated in a weekly dance program, compared to a Reference group who remained physically inactive.

**Methods:** This six-year longitudinal observational study included 43 PwPD who attended weekly dance classes and were evaluated using the Mini-Mental State Examination (MMSE) and Movement Disorder Society–Unified Parkinson’s Disease Rating Scale (MDS-UPDRS). A Reference group of 28 PwPD, matched on age, gender, and Hoehn & Yahr scores, were selected from the Parkinson’s Progression Marker Initiative, and assessed using the MDS-UPDRS and Montreal Cognitive Assessment (MoCA). Cognitive scores were standardized. Generalized estimating equations were used to compare cognitive and gait outcomes across time.

**Results:** The Dance group was significantly different from the Reference group (*p* < 0.001), with improved cognitive scores in 2016, 2017, and 2018. The Dance group had worse gait at baseline, however, the Reference group showed significantly poorer gait performance by 2018. No significant association was found between gait and cognitive scores.

**Conclusion:** After two years of weekly dance, the Dance group showed improvements in cognition and maintained stability in gait performance. The findings highlight the potential neuroprotective benefits of continued dance engagement over six years.

## Introduction

Parkinson’s Disease (PD) and Alzheimer’s Disease (AD) are regarded as the most prevalent age-related neurodegenerative disorders, affecting millions around the world.^1,2^ Although the pathophysiology of both conditions differs, PD and AD may share overlapping neuropathological features.^3^ AD is defined by the progressive loss of cognitive functions, including memory and learning^1^, while PD, initially documented by James Parkinson as “Shaking Palsy” in 1817^4^, is now primarily characterized as a movement disorder with hallmark motor symptoms, including bradykinesia, tremors, and rigidity.^5^ Similar to AD, the prevalence of PD is also age-related, impacting approximately 1% of persons between the ages of 65 and 69 years, increasing to approximately 3% for individuals at 80 years of age and older.^6,7^ In the absence of a cure, the prevalence and societal burden of PD is expected to increase as the global population continues to age, signifying the need for therapeutic interventions.^5,8,9^

Aside from the classic motor signs of PD, persons with PD (PwPD) may suffer from additional motor and non-motor symptoms.^10^ Gait disturbances are among the most debilitating motor symptoms of PD, affecting nearly all individuals as the disease progresses.^11,12^ Greater fall risks are associated with impaired gait, which contributes to the need for residential care and loss of independence.^13^ Furthermore, non-motor symptoms, particularly cognitive impairment, including mild cognitive impairment (MCI) and dementia, are relatively common in PD. A decline in cognition is regarded as a predictor of reduced quality of life and increased morbidity as well.^3,14^ Gait and cognitive dysfunction often overlap, especially in dual-task situations requiring attention and executive function, which exacerbates the need for interventions that address both motor and cognitive domains simultaneously.^15,16^ Recently, Zhou et al^17^ found that dopaminergic dysfunction in the prefrontal cortex may contribute to the cognitive-motor dual-task impairments that are seen among PwPD. Furthermore, advances in neuroimaging, including studies that have employed resting-state functional MRI, have also enhanced our understanding of the neural mechanisms driving cognitive decline in PwPD.^18^

As researchers continue to investigate ways to support brain health in PwPD, Dhami, Moreno, and DeSouza^19^ proposed a framework in 2014 emphasizing dance as a multimodal form of neurorehabilitation that combines physical and cognitive stimulation with music and emotional engagement. Dance, which also integrates rhythm and social engagement, has become a promising intervention for PwPD, with numerous studies demonstrating improvements in motor and non-motor symptoms of PD, including cognitive and gait functioning.^20,21^ Dance is associated with neural plasticity, which may lead to improved motor planning and execution.^22^ Recent findings have demonstrated consistent improvements in gait variability, stride length, and dual-task performance among PwPD after dance engagement, which highlight the neuroprotective benefits of dance.^23^ Moreover, Fiorenzato et al^24^ explored the complexity of brain dynamics in PD as a marker of cognitive decline, offering a framework for exploring the effect of dance on neural reorganization and cognitive resilience. Studies have shown dance may lead to improved cognition, particularly among the executive, attention, and memory domains.^20,25^ A recent study by Chen et al^26^ also highlights the relevance of spontaneous brain activity in hippocampal areas as a potential biomarker for cognitive dysfunction, providing new avenues to examine how dance can improve cognitive outcomes by altering these neural processes.

Further research has also shown improvements among additional non-motor symptoms, such as anxiety and depression, in PwPD after dance engagement.^23,27^ Through creativity and artistry, the Dance for Parkinson’s Disease program may promote physical, emotional, and social engagement, which extend beyond the limitations and challenges of the disease.^28^ With these encouraging findings we examine the long-term impact of dance on motor and cognitive abilities is essential to optimize therapeutic interventions and ultimately improve quality of life in PwPD.

## Methods

### Study participants

Volunteers were recruited from Canada’s National Ballet School and Trinity St. Paul’s Church in Toronto, Ontario, Canada. The study sample included a total of 43 PwPD across the six-year study period from 2014 to 2019 (Dance group; *N_male_* = 25; *M_age_* = 70.0, *SD* = 7.0), with mild severity (*M_H&Y_* = 1.3, *SD* = 0.8). All 43 PwPD in the Dance group underwent a minimum of one data collection session between 2014 to 2019. However, the gait analysis was limited to a subsample of *n* = 10 people who completed the Movement Disorders Society–Unified Parkinson’s Disease Rating Scale (MDS-UPDRS) at least twice. The study was approved by the Office of Research Ethics Committee (ORE) of York University (approval numbers 2013-211 & 2017-296) on November 07, 2013 & September 29, 2017. Written informed consent was obtained from all volunteers in accordance with the approved protocol.

A Reference group of PD-non-dancers was selected from a larger cohort of persons with PD from the Parkinson’s Progression Marker Initiative (PPMI), a longitudinal study aimed at identifying biomarkers for PD and funded by the Michael J. Fox Foundation (MJFF). Matching was prioritized to ensure comparability between groups, and a total of 28 people with PD were chosen for the Reference group and were matched on key factors such as age (*M_age_* = 66.7, *SD* = 7.3), gender (*N_male_* = 21) and scores on the H&Y scale (*M_H&Y_* = 1.4, *SD* = 0.8). The Reference group was further selected based on the Leisure Time Activity subscale of the Physical Activity Scale for the Elderly (PASE). The sub-scale was used to identify subjects based on their engagement in physical activities, including aerobic or ballroom dance. No people with PD in the Reference group engaged in any form of physical exercise throughout the duration of the present study.

### Measures

Baseline cognitive and motor data for the Dance group were collected from each volunteer regardless of the number of dance classes attended. The Mini-Mental State Examination (MMSE) was administered before each dance session. Before each dance session, part III of the Movement Disorder Society–Unified Parkinson’s Disease Rating Scale (MDS-UPDRS), which assesses motor aspects of daily living, was collected, with gait being specifically scored using item 3.10 of the motor examination. At the end of each session, the remaining parts I, II, and IV, which assessed non-motor symptoms, aspects of daily living, and complications, were evaluated. Item 3.10, labelled gait, of the UPDRS was scored by four raters, all of whom were trained following the completion of The International Parkinson and Movement Disorder Society online certificate training program.

Cognitive and motor data for the Reference group were obtained from the PPMI database for the years 2014 to 2019. Cognitive scores were assessed using the Montreal Cognitive Assessment (MoCA), and motor scores were collected using the UPDRS.

### Procedure

The Dance group participated in 75-min weekly dance classes based on the Dance for Parkinson’s program^29^ and were led by two trained dance instructors. These classes were offered as part of the Sharing Dance Parkinson’s program at Canada’s National Ballet School and the Dancing with Parkinson’s program at Trinity St. Paul’s Church, both located in Toronto, Ontario, Canada. Each class began with a seated warm-up, followed by “barre” exercises, and sessions ended with floor-work dances. The Dance group also learned a specific choreography in preparation for an upcoming performance (see Bearss et al., 2017 for the full dance protocol) during the first years.^30^

### Statistical Analyses

To allow for a fair comparison of cognitive scores between both groups, raw MMSE scores (Dance group) and MoCA scores (Reference group) were standardized into z-scores within each group. Standardization was performed using the formula: *z* = (*x* – *M*) / *SD*, where *x* is the participant’s raw cognitive score, *M* is the group mean, and *SD* is the group standard deviation. This approach was the preferred method for comparing scores from different cognitive tests as it has been applied in previous research.^31,32^ A generalized estimating equations (GEE) model allowed for group comparisons between cognitive scores, gait scores, and the relationship between cognitive and gait scores across time, while accounting for repeated measures across all participants in both groups. The data analyses and statistics were performed using Python (JupyterLab, version 3.6.3).

## Results

### Descriptive Statistics

The mean MMSE scores for the Dance group were 28.34 (*SD* = 2.10), as shown in Figure 1, ranging from 21.0 to 30.0. The number of PwPD with MMSE scores per year were: 2014 (*n* = 10), 2015 (*n* = 9), 2016 (*n* = 23), 2017 (*n* = 12), 2018 (*n* = 24) and 2019 (*n* = 5). In the Reference group, the mean MoCA score was 25.86 (*SD* = 2.83), with scores ranging from 16.0 to 30.0, as shown in Figure 2. The number of people with PD with MoCA scores per year were: 2014 (*n* = 14), 2015 (*n* = 17), 2016 (*n* = 18), 2017 (*n* = 21), 2018 (*n* = 18), and 2019 (*n* = 18). Figure 3 shows the mean standardized cognitive scores for the Dance group were 0.82 (*SD* = 0.23), and 0.70 (*SD* = 0.20) for the Reference group. For gait scores, the Dance group had a mean of 0.7 (*SD* = 0.4), ranging from 0.0 to 1.25. The Reference group had a mean of 0.6 (*SD* = 0.6), ranging from 0.0 to 3.00 for gait scores.

**Figure 1.**
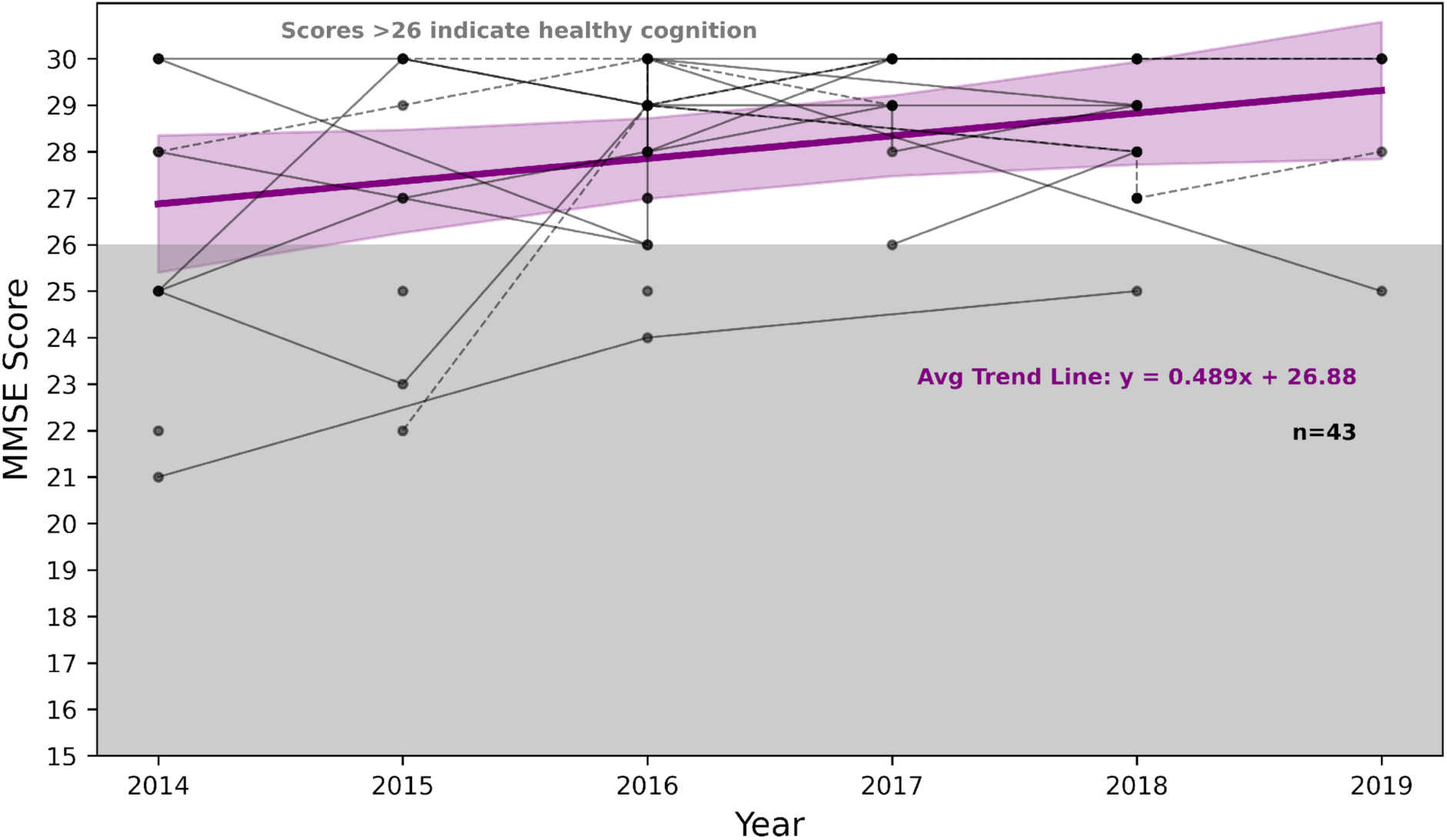
MMSE scores for the Dance group (*n* = 43) across time. The number of people with PD with MMSE scores per year were: 2014 (*n* = 10), 2015 (*n* = 9), 2016 (*n* = 23), 2017 (*n* = 12), 2018 (*n* = 24) and 2019 (*n* = 5). The bold purple line shows the average trend in MMSE scores across time, with the surrounded shaded area representing the 95% confidence interval. Points within the shaded grey area indicate cognitive impairment. Solid lines represent male people with PD, dashed lines represent female PwPD.

**Figure 2.**
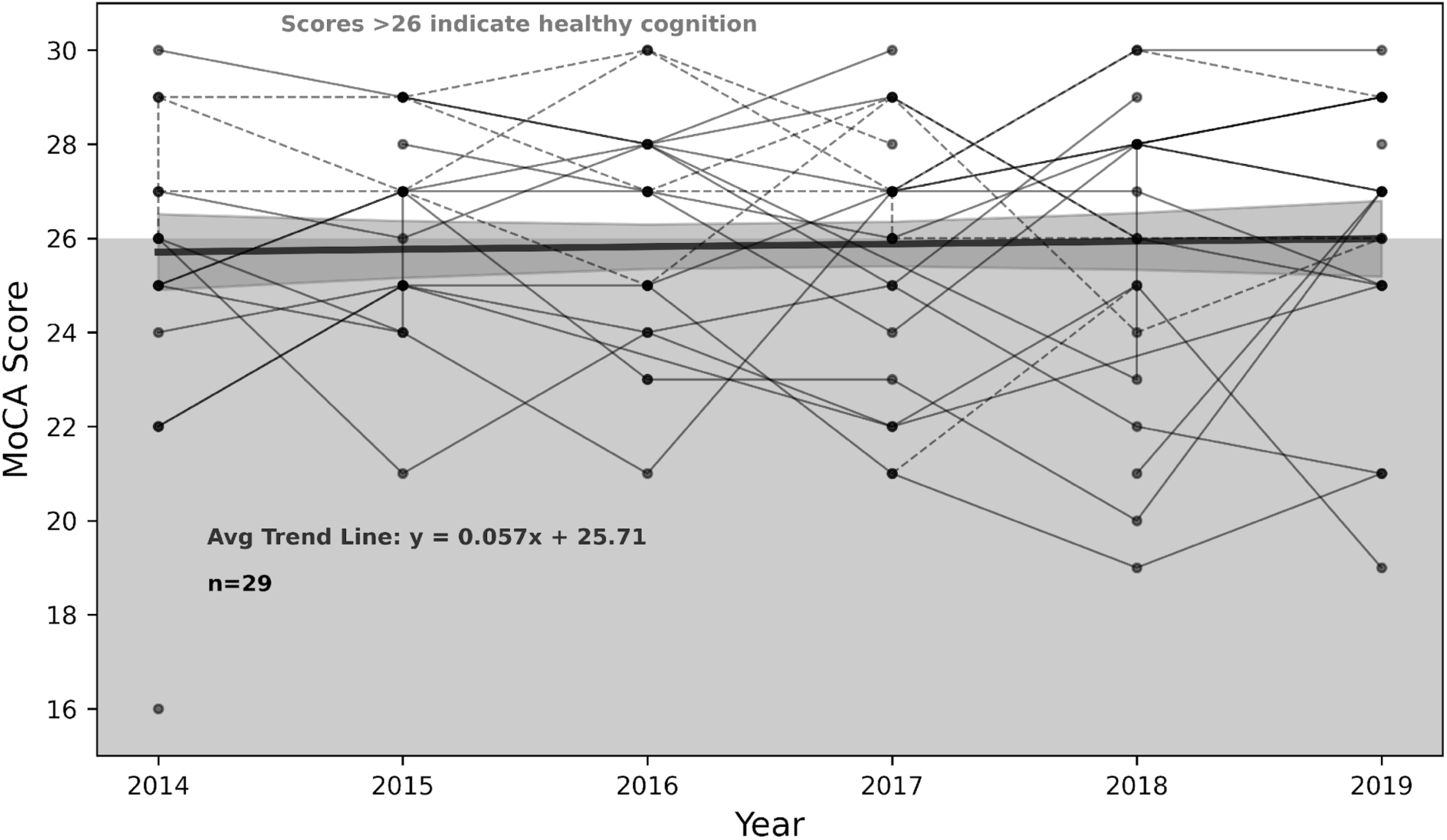
MoCA scores for the Reference group (*n* = 28) across time. The number of people with PD with MoCA scores per year were: 2014 (*n* = 14), 2015 (*n* = 17), 2016 (*n* = 18), 2017 (*n* = 21), 2018 (*n* = 18), and 2019 (*n* = 18). The bolded black line shows the average trend in MoCA scores across time, with the surrounded shaded area representing the 95% confidence interval. Points within the shaded grey area indicate cognitive impairment. Solid black lines represent male PwPD, while dashed lines represent female PwPD.

**Figure 3.**
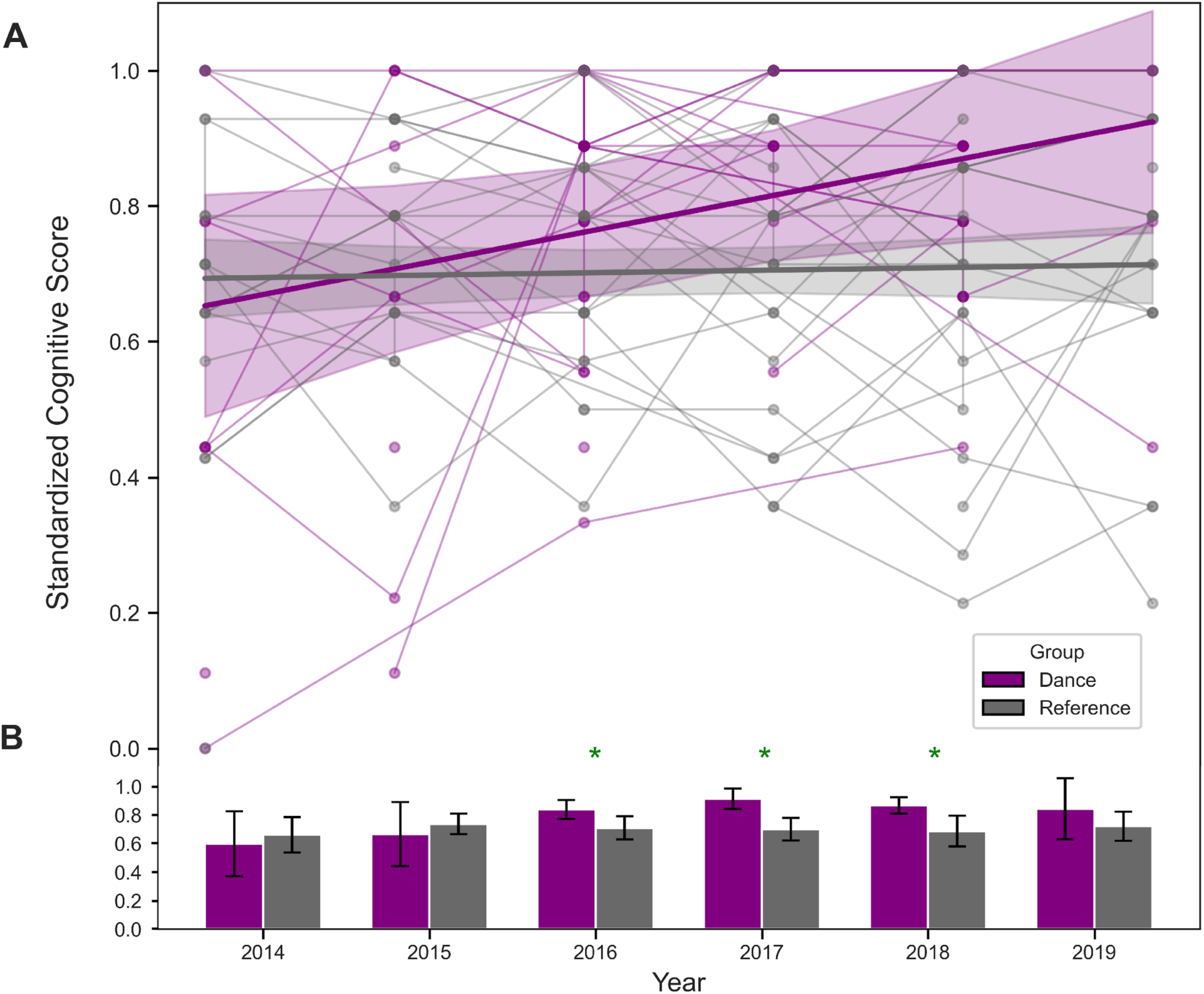
**A:** Standardized cognitive scores for the Dance group (purple) and Reference group (grey) plotted individually across time (from Fig 1 and 2). The bolded purple and grey lines represent the average trends in standardized cognitive scores for the Dance and Reference groups, respectively, across time. Shaded regions surrounding each line represent the 95% confidence intervals. **B:** Group-level standardized scores for the Dance and Reference group, showing significant cognitive improvements for the Dance group in 2016, 2017, and 2018 compared to the Reference group. An asterisk signifies a significant difference between group means. Error bars represent the 95% confidence intervals.

### Cognitive Score Differences Over Time

Generalized Estimating Equations (GEE) revealed a significant difference in cognitive scores between the Dance and Reference groups over time, with the Dance group demonstrating higher scores overall (*p* < 0.001). As shown in Figure 3, cognitive scores in the Dance group were significantly higher in 2016 (β = 0.2391, *p* = 0.028), 2017 (β = 0.3145, *p* = 0.007), and 2018 (β = 0.2675, *p* = 0.010), although this improvement was no longer significant in 2019 (β = 0.2444, *p* = 0.096). Figures 1-3 further illustrate that the Dance group showed greater improvement over the six years compared to the Reference group.

### Group and Time Effects on Cognition

The Group x Year interaction terms from the GEE model found no significant cognitive score differences between the Dance and Reference groups from 2014 to 2015 (β = 0.0095, *p* = 0.950), as shown in Figure 3. In 2016, the Reference group had a trend towards lower cognitive scores compared to the Dance group, but this was not statistically significant (β = -0.1907, *p* = 0.134). In 2017, cognitive scores of the Reference group were significantly lower than the Dance group (β = -0.2751, *p* = 0.036). This trend continued in 2018 with the Reference group scoring lower than the Dance group, although this difference was marginally significant (β = -0.2415, *p* = 0.058). No significant difference was found between the two groups in 2019 (β = -0.1841, *p* = 0.258).

To further explore group differences at each time point, Mann-Whitney U tests with Bonferroni correction were conducted. These post hoc comparisons assessed differences between both the Dance and Reference groups within each year and not changes relative to baseline. No significant differences were found at our 2014 baseline (*p* = 0.977) and 2015 (*p* = 0.938). Most importantly, the Dance group scored significantly higher than the Reference group in 2016 (*p* = 0.008), 2017 (*p* = 0.001), and 2018 (*p* = 0.0042). No significant group differences were found in the last year of 2019 (*p* = 0.1758).

### Analysis of Gait Over Time

A GEE model was performed to assess the effects of group and year on gait performance. The model included main effects for group, year, and their interaction. An independent correlation structure was used to account for repeated measures. The results showed that the intercept was significantly different from zero (β = 0.8571, *p* < 0.001), suggesting that the Dance group had worse gait performance at baseline (2014). This suggests there were baseline differences in gait between both groups. As shown in Figure 4, the Reference group had lower gait scores, indicating better gait performance at baseline, compared to the Dance group. The GEE analysis confirmed this difference was statistically significant (β = −0.3665, *p* = 0.012). In 2018, the Reference group showed significantly worse gait scores compared to the Dance group (β = 0.4946, *p* = 0.007), although no other group differences reached significance.

**Figure 4.**
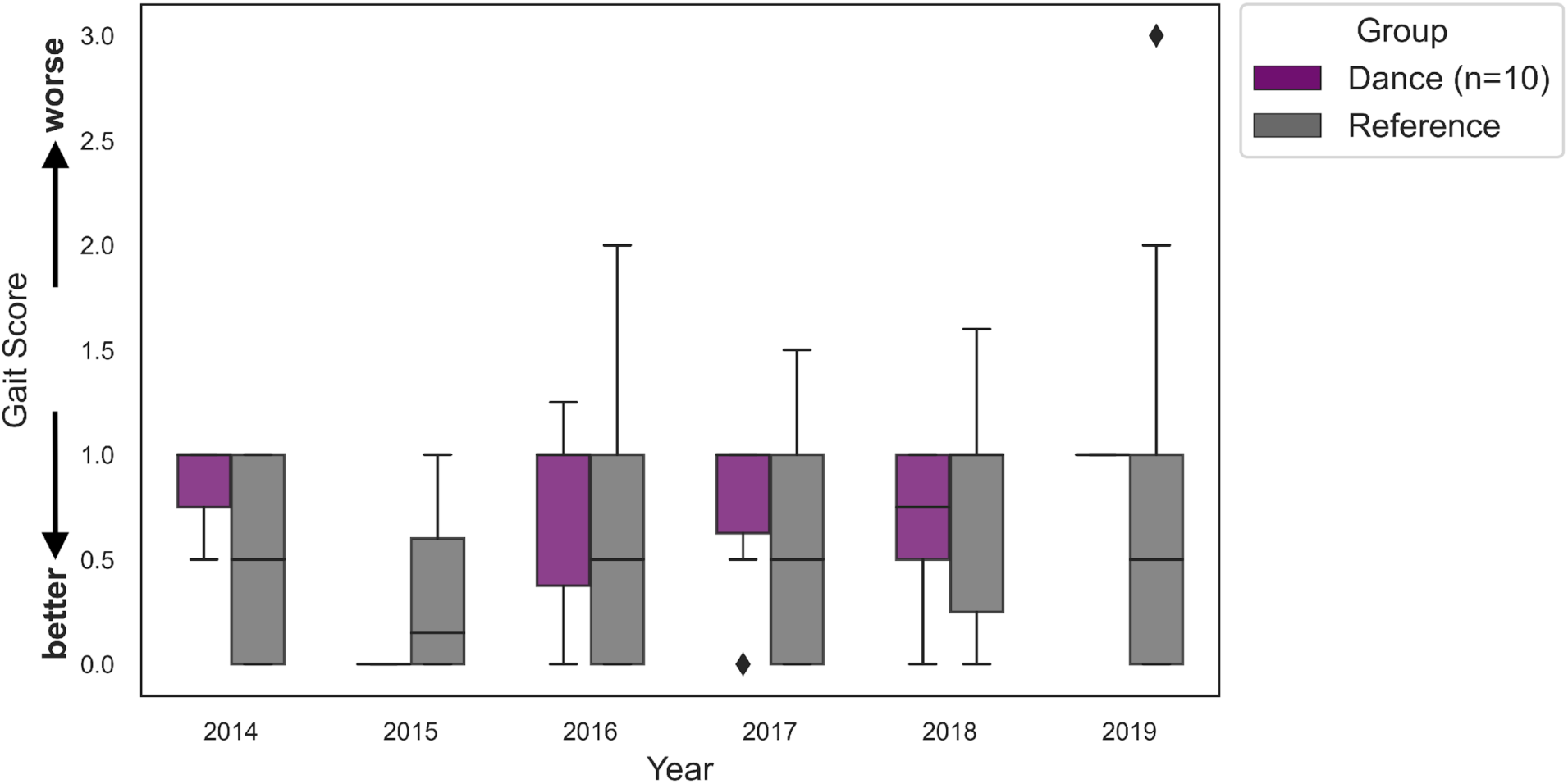
Gait scores over time for the Dance group (*n* = 10, purple) and Reference group (grey). Higher scores indicate worse gait performance. Boxplots represent the distribution of gait scores for each group from 2014 – 2019.

### Group and Gait Effects on Cognition

A separate GEE model was conducted to examine cognition as a function of gait, group, and year. The results showed that the intercept for cognition was significantly different from zero (β = 0.7392, *p* < 0.001), thus indicating the Dance group had better cognition at baseline. There was no significant main effect of group (β = -0.0524, *p* = 0.630), and no significant association was found between gait performance and cognition (β = -0.0476, *p* = 0.307), suggesting that gait performance was not directly associated with cognitive function.

## Discussion

Our findings demonstrate that long-term dance participation was associated with improved cognitive outcomes in PwPD. At baseline (2014) and in 2015, there were no significant differences observed between both groups. From 2016 to 2018, cognitive scores of the Dance group were significantly higher compared to those of the Reference group. However, this significance was no longer meaningful by 2019, likely attributed to a decline in participation in the Dance group, leading to reduced statistical power. The upward trend observed in the Dance group demonstrates the potential cognitive benefits of dance for PwPD, which is consistent with previous literature.^32^ McKee and Hackney^33^ conducted a 12-week adapted tango intervention that showed improvements in spatial cognition and executive functioning in PwPD, as measured by the MoCA, Reverse Corsi Blocks, and Brooks Spatial Task. Similarly, Michels et al.^34^ reported a mean increase of 0.44 in MoCA scores in PwPD following a 10-week dance intervention, compared to a mean decrease of 0.50 in the control group, although the study did not test for statistical significance. Duarte et al.^35^ also reported significant improvements in executive functioning among PwPD following a six-month dance-based intervention, as measured by the Frontal Assessment Battery. Given that most dance-based studies assessing cognitive function in PwPD are relatively short^32^, the present study provides a better understanding of the long-term cognitive impact of dance, with findings suggesting preservation of global cognition among PwPD over a six-year period.

These findings may translate to AD and other neurodegenerative conditions as well. Through motor learning, dance engages certain areas of the brain involved in memory and executive functioning, which may promote neural resilience.^36^ Previous research has shown that dance-based interventions led to improved cognitive outcomes, specifically within memory, executive functioning, and other cognitive domains among persons with mild cognitive impairment, AD, and dementia.^37,38^ Additional research has further demonstrated that dance may lead to improved mood and quality of life among persons with AD^39^, and gait speed among persons with dementia.^40^ Abreu and Hartley^41^ also found significant improvements in functional ability in a case study involving an individual with AD and other comorbidities.

As demonstrated by Bearss & DeSouza^42^, dance may help to maintain mobility in PwPD. While gait performance in the Dance group was relatively stable across time, our results should be interpreted with caution. A post-hoc power analysis revealed that with our sample of 10 volunteers who had both gait and cognitive measurements, the study had only 3.3% power to detect the observed effect size (β = -0.0377), indicating that the analysis was severely underpowered to detect potentially meaningful associations between these variables. In comparison to the Dance group, the Reference group (*n* = 28) showed a significant decline in 2018. Given that dance involves complex movements and postural control^43^, it is possible that improvements in gait performance within the Dance group may not have been fully captured by the single item gait measure (UPDRS item 3.10) used in the present study. Additionally, the small sample size for this gait analysis overlapped with volunteers from our Bearss & DeSouza^42^ study, which included a larger sample from the Dance group (*n* = 16) and assessed motor symptoms using the complete UPDRS Part III motor examination.

Despite the literature suggesting a relationship between gait and cognition in PD^44–46^, our analysis did not find a significant association between these two variables because of this small subset of 10 PwPD. Previous literature by Morris et al^47^ found that specific aspects of gait, mainly pace and turning, are linked to distinct cognitive domains, including attention, executive functioning, and memory. Further research has also demonstrated a relationship between specific aspects of gait, such as gait speed, and distinct cognitive domains, particularly executive dysfunction.^48,49^ In contrast, our study assessed global cognition, rather than individual cognitive domains, and analyzed gait as a single measure in a subset of our population. In addition, the lack of a significant association between gait and cognition in our study may be attributed to the limited number of gait assessments available for analysis, as well as the reduced sample size.

### Strengths and Limitations

To our knowledge, this is the first study to assess the long-term effects of dance on cognition among PwPD across a six-year period. The longitudinal design of the present observational study provides valuable insights into the benefits of long-term dance engagement as a non-pharmacological intervention, in addition to standard treatment practices. Additionally, the use of a community-based dance program highlights real-world applicability which may help to refine future research methodologies, however, there are a few limitations that should be acknowledged in our present study. Participation in the Dance group was non-restrictive, resulting in variability across PwPD. As a result, PwPD with lower engagement may not have experienced the same level of cognitive benefits as those with higher engagement. In addition, gait assessments were available for only 10 PwPD in the Dance group, as only those with a minimum of two testing sessions were included in the gait analysis to assess pre- and post-changes in gait performance, limiting the statistical power for detecting changes in gait across time.

Lastly, the type of cognitive assessments used differed between the two groups. The Dance group was assessed using the MMSE while MoCA was administered in the Reference group. Although both assessments were standardized, the MMSE has shown reduced sensitivity to cognitive changes in PwPD when compared to the MoCA.^49–52^ However, the MMSE has demonstrated good test-retest reliability when assessed among older individuals with normal cognition.^53^ Truong et al^54^ found that MMSE scores among cognitively healthy older adults remained consistent with memory performance throughout a six-year period. While older adults with dementia may show retest effects, as demonstrated by Gross et al.^55^, these effects are substantially smaller when compared to older adults with normal cognition. This suggests that cognitive impairments may limit participants’ ability to benefit from familiarity with the assessment tool.^51^ Although retest effects using the MMSE among PwPD have not been explored, the difference in measurement sensitivity between the MoCA and MMSE may have influenced our findings. As the Reference group, assessed using the MoCA, showed a trend towards poorer cognitive performance over time, it is possible that similar changes in the Dance group may not have been fully captured due to the limitations mentioned above of the MMSE.

Given the range of cognitive problems experienced by PwPD, future research should further investigate the effectiveness of dance for improved cognitive functioning using more sensitive cognitive assessments for PwPD, using attention, language, working memory tasks and social cognition with incorporating neuroimaging and biomarkers to assess neural mechanisms of cognitive changes. To better understand the relationship between cognitive and gait outcomes following long-term dance engagement, future research should utilize more comprehensive gait assessments as well.

## Data Availability

Data is available on request in accordance with the ethics used at the time of data collection.

## Acknowledgements

Thanks to all the people from our lab over the years for the project and all the people at our testing locations. We are indebted to S. Leung, R. Cohan, P. Dhami, S. Maguire, D. Rabinovich, H. Tehrani, K McDonald, and R. Andrew for invaluable assistance during all phases of the research program. We are also thankful for the collaborations with Canada’s National Ballet School (A. Seto, A. Powell) and Trinity St. Paul’s Church in Toronto, ON, Canada, and D. Leventhal from Dance for Parkinson’s at Mark Morris Dance Group for continued training and support.

## Author Contributions

Simran Rooprai (Formal Analysis; Visualization; Writing – Original Draft); Harsimran Dogra (Formal Analysis; Writing – Original Draft); Ashkan Karimi (Formal Analysis; Visualization); Rafia Rafique (Writing – Original Draft); Emily D’Alessandro (Data curation); Karolina Bearss (Conceptualization; Methodology; Investigation; Project Administration; Writing – Review & Editing); Sarah Robichaud (Resources; Project Administration); Rachel J. Bar (Resources; Project Administration); Joseph F.X. DeSouza (Conceptualization; Methodology; Investigation; Formal Analysis; Resources; Project Administration; Supervision; Funding acquisition; Writing – Review & Editing).

## Statements and Declarations

### Ethical considerations

This study was approved by the Office of Research Ethics Committee (ORE) of York University (approval no. 2013-211 & 2017-296) on November 07, 2013 & September 29, 2017.

### Consent to participate

Written informed consent was obtained from all volunteers.

### Consent for publication

Not Applicable.

### Data availability

Data is available on request in accordance with the ethics used at the time of data collection.

